# Time to cardiovascular magnetic resonance imaging influences diagnostic yield in patients with suspected myocardial infarction with nonobstructive coronary arteries: a meta-analysis

**DOI:** 10.1101/2025.03.03.25323213

**Authors:** Guy D Eslick, Enid M Eslick, Martin Ugander, Rebecca Kozor

**Author notes:** Address for Correspondence: Guy D. Eslick, PhD, The University of Sydney, Australia. MU and RK contributed equally to this work.

## Abstract

**Objectives:** To conduct a systematic review and meta-analysis to assess the evidence of cardiovascular magnetic resonance imaging (CMR) in patients with suspected myocardial infarction and nonobstructive coronary arteries (MINOCA), and how time to CMR influences diagnosis.

**Background:** CMR is indicated in patients with suspected MINCOA but it is unclear when is best to perform the CMR and how this timing can influence diagnosis.

**Methods:** We systematically conducted a comprehensive literature search to identify relevant studies. These studies were assessed to determine the study quality and analysis was performed using a Random-effects model.

**Results:** There were 23 eligible studies, including 4,231 patients. The mean quality score was 9.35 out of 10. For MINOCA patients assessed by CMR, the average median time from presentation to CMR was 12.50 days (SD: 14 days, range 0-365 days). The pooled frequencies of the most common diagnoses were: myocarditis (29%), myocardial infarction (22%), Takotsubo syndrome (10%), cardiomyopathy (7%), and 22% had no cardiac diagnosis (normal CMR findings). In pooled analysis, the prevalence of normal CMR findings increased by three percentage points for each extra day of waiting between presentation and CMR scanning over the studied range of 0-14 days (Slope: 3.1 %-points/day; r=0.67, p=0.003).

**Conclusions:** In patients with suspected MINOCA, the longer it takes a patient to have a CMR scan, the more likely the results will be normal and no diagnosis made by CMR. CMR should be performed as early as possible in suspected MINOCA.

**Highlights:**

- In patients with suspected MINOCA, the pooled frequency of diagnoses made by CMR include myocarditis (29%), myocardial infarction (22%), Takotsubo syndrome (10%), cardiomyopathy (7%), and no diagnosis/normal study (22%).
- The longer it takes to have a CMR scan the more likely the patient will have a normal scan.
- In patients with suspected MINOCA, CMR scanning should be performed as soon as possible.

## INTRODUCTION

Myocardial infarction with non-obstructive coronary arteries (MINOCA) is the term used for patients presenting with symptoms suggestive of myocardial ischaemia, cardiac enzyme elevation, and electrocardiographic (ECG) changes, but do not have obstructive coronary artery disease (>50% stenosis) on angiography (1). MINOCA occurs in up to 20% of patients with acute ST-segment elevation myocardial infarction (STEMI) or non-ST-segment elevation myocardial infarction (NSTEMI) (2). There are multiple potential underlying diagnoses in patients presenting with a working diagnosis of MINOCA including but not limited to acute myocarditis, Takotsubo syndrome, other cardiomyopathies, and myocardial infarction that is due to coronary artery vasospasm, embolism, dissection, microvascular dysfunction, or plaque disruption that is non-obstructive (3). This latter group is considered ’true’ MINOCA. The correct diagnosis in suspected MINOCA is important to identify because this can affect subsequent management and outcomes.

Cardiovascular magnetic resonance (CMR) is a powerful non-invasive imaging modality that can play a pivotal role in the diagnosis of some of these differential diagnoses and has been shown to provide an accurate diagnosis in 65-90% of suspected MINOCA cases (4, 5). Studies have shown that using CMR in the management of unexplained chest pain patients in the emergency department is safe, feasible, and cost-effective compared to traditional management approaches of chest pain patients (6, 7). Due to this, CMR has been recognised internationally as an appropriate diagnostic tool in suspected MINOCA and the use of CMR in the assessment of suspected MINOCA is recommended by the American Heart Association (AHA) and the European Society of Cardiology (ESC) (8, 9). However, the best timing for performing CMR in relation to presentation is unknown. Current practices may be *ad hoc* and/or dependent on the physician, patient and health system in question.

We therefore conducted a systematic review and meta-analysis to assess the existing evidence of CMR in patients with a working diagnosis of MINOCA (suspected MINOCA) in order to determine (a) the relationship between time to CMR from presentation and the diagnosis made by CMR, and (b) the frequency of underlying causes as identified by CMR.

## METHODS

### Study Protocol

We followed the Preferred Reporting Items for Systematic reviews and Meta-Analyses (PRISMA) guidelines (10). A systematic search of the databases MEDLINE, EMBASE, and Google Scholar was performed up to 24 September, 2024, to identify relevant articles (Figure 1). Our search items included “myocardial infarction with nonobstructive coronary arteries” OR “MINOCA” AND “cardiovascular magnetic resonance imaging” OR “CMR”. The reference lists of relevant articles were also searched for appropriate studies. No language restrictions were used in either the search or study selection. No search for unpublished literature was performed. Approval by an ethics committee for this study was not applicable since it is a meta-analysis. All data included in the present study are available upon reasonable request to the authors.

**Figure 1.**
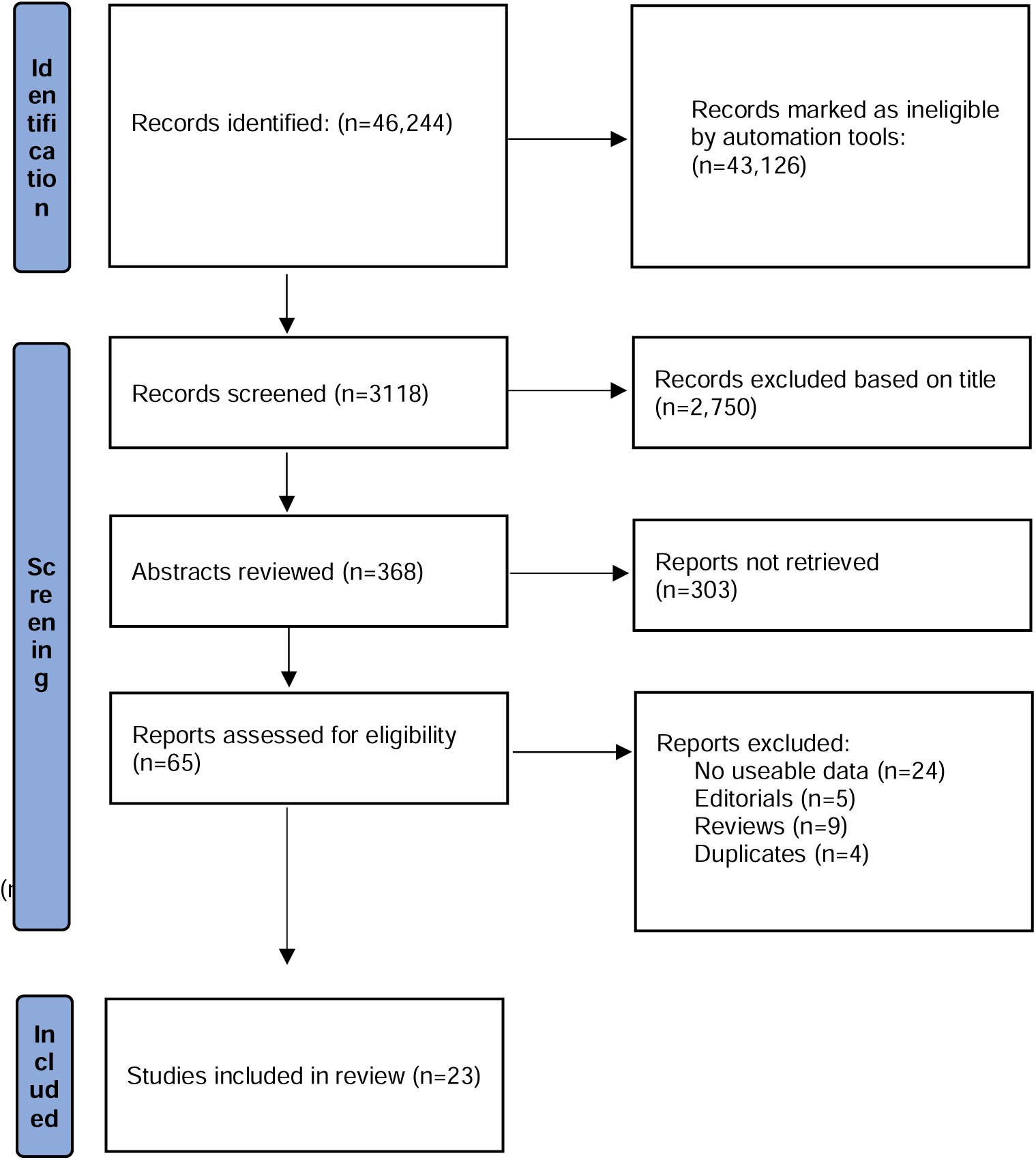
Flowchart of the search strategy.

### Study Selection

We included studies that met the following inclusion criteria: (1) studies that assessed suspected MINOCA patients using CMR; (2) studies where the prevalence estimate was reported for various outcomes; (3) the 95% confidence interval (CI) was reported, or the data were presented such that the CI could be calculated. We excluded studies that did not meet these inclusion criteria.

### Data Extraction

The data extraction was performed using a standardised form, collecting information on the prevalence of conditions based on CMR imaging, publication year, study design, population type, country, sample size, percent female, and mean age. Quality of the studies was assessed using the JBI Checklist for Case Series, the instrument (11).

### Statistical Analysis

Pooled prevalence estimates and 95% confidence intervals were calculated for the various cardiac conditions diagnosed by CMR imaging in MINOCA patients, using a random-effects model (12). Heterogeneity was assessed using the I² statistic with results of 0%-25% (low), 25%-75% (moderate), and >75% (high) levels of heterogeneity (13). We assessed publication bias using the Egger’s regression model only if there were greater than ten studies (14). All analyses were performed with Comprehensive Meta-Analysis, Version 4. Borenstein, M., Hedges, L., Higgins, J., & Rothstein, H. Biostat, Englewood, NJ (2022). Correlations were assessed using Pearson correlation coefficient (r) for time to CMR in days with the different diagnoses using GraphPad Prism v9.0.

## RESULTS

### Search strategy

The literature search strategy initially identified 46,244 potential articles (Figure 1). Papers were excluding based on searches of title and abstract before reading full-text of selected articles.

### Study characteristics

Overall, there were 23 studies that met the inclusion criteria and were included in the meta-analysis (15–38). These studies consisted of 4,231 patients with a mean age of 53 years (SD: 5.29 years, range: 44-62 years), and 48% were female.

### Quality assessment

The mean quality score was 9.35 (range 8 to 10). Of the 23 studies included, 11 studies scored ten.

### Time to CMR

For patients with suspected MINOCA, the average median time from presentation to CMR imaging was 12.50 days (SD: 14 days, range 0-365 days). The time to CMR varied by geographic region: in one multinational study (Australia, Europe, United States) the average median time was 7 days (n=1), in Australia the average median time was 10 days (n=1), in the United States the average median time was 4.3 days (n=3), in Europe the average median time was 7 days (n=13), in the United Kingdom the average median time was 29 days (n=5).

### Diagnosis by CMR

#### Myocarditis

The pooled prevalence of myocarditis was 29.0% (95% CI: 20.0%-39.0%; I^2^=96.34, p<0.0001), amongst those with suspected MINOCA who underwent CMR. There was no correlation between time to CMR and the diagnosis myocarditis (r=-0.16, p=0.54).

#### Takotsubo syndrome

The pooled prevalence of Takotsubo syndrome was 10.0% (95% CI: 7.0%-14.0%; I^2^=89.60, p<0.0001), amongst those with suspected MINOCA who underwent CMR. There was no correlation between time to CMR and the diagnosis Takotsubo cardiomyopathy (r=-0.32, p=0.24).

#### Myocardial infarction

The pooled prevalence of myocardial infarction was 22.0% (95% CI: 17.0%-27.0%; I^2^=88.90, p<0.0001), amongst those with suspected MINOCA who underwent CMR. There was no correlation between time to CMR and the diagnosis myocardial infarction (r=-0.33, p=0.16).

#### Cardiomyopathy

The pooled prevalence of cardiomyopathy was 7.0% (95% CI: 4.0%-13.0%; I^2^=95.96, p<0.0001), amongst those with suspected MINOCA who underwent CMR. There was a moderate negative correlation between time to CMR and the diagnosis cardiomyopathy (r=-0.62, p=0.04).

#### No cardiac diagnosis on CMR

The pooled prevalence of no cardiac diagnosis found on CMR (ie, a normal CMR study) was 22.0% (95% CI: 16.0%-29.0%; I^2^=92.32, p<0.0001), amongst those MINOCA patients who had CMR. There was a moderate positive correlation between time to CMR imaging and a normal CMR study result (r=0.67, p=0.003) (Figure 2). We conducted additional sub-group analyses where time to CMR outliers (above 14 days) were removed and the correlation strengthened (r=0.73, p=0.001). Over the median range of 3-14 days after presentation, the regression line was described by the function (CMR normal findings [%]) = 3.05 x (days between presentation and CMR) + 4.58. This shows that the fraction of normal findings by CMR increases by three percentage points for each additional day of waiting between presentation and CMR scanning.

**Figure 2.**
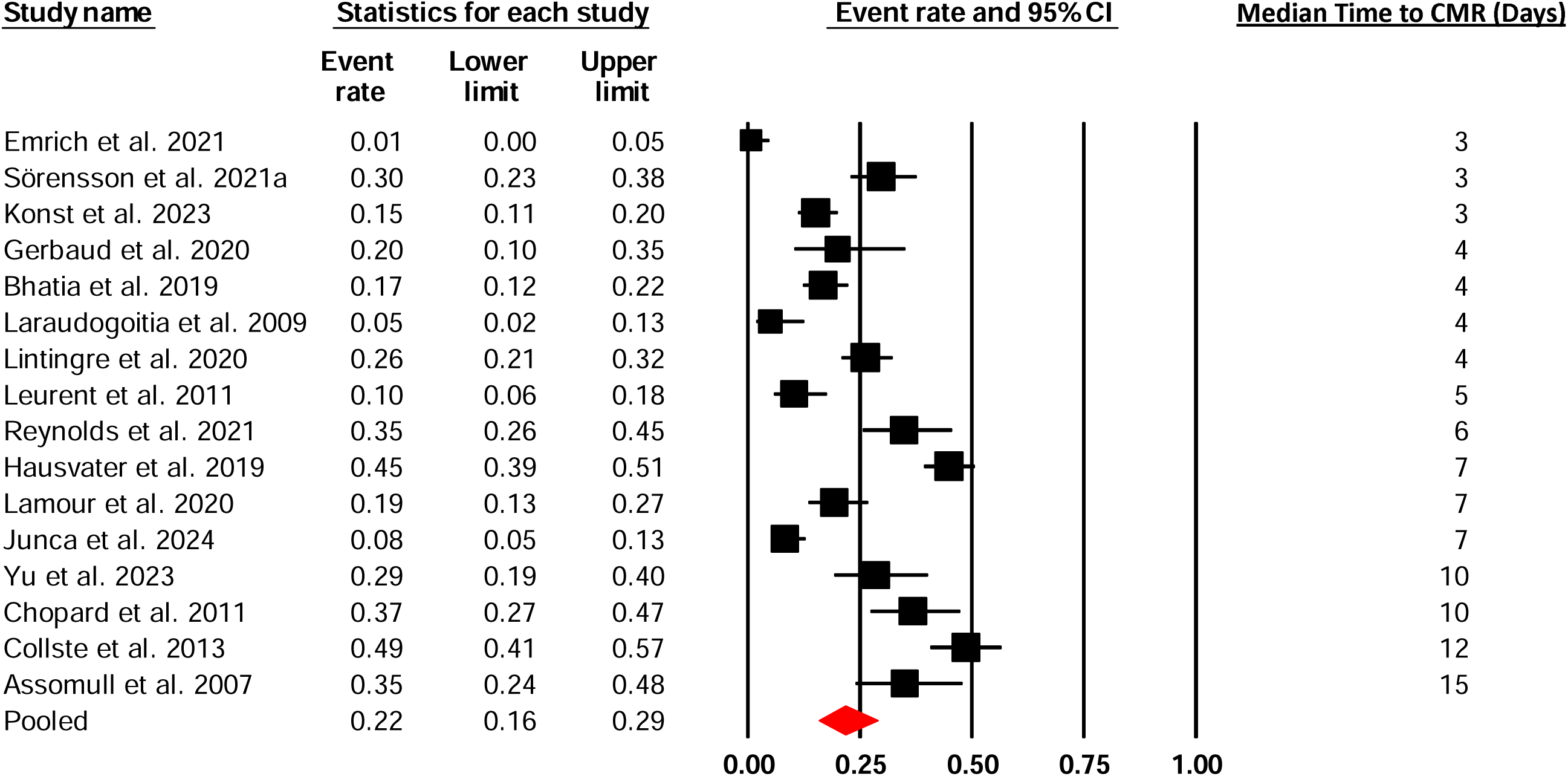
Forest plot of the prevalence of normal CMR findings among those with MINOCA who had a CMR.

**Figure 3.**
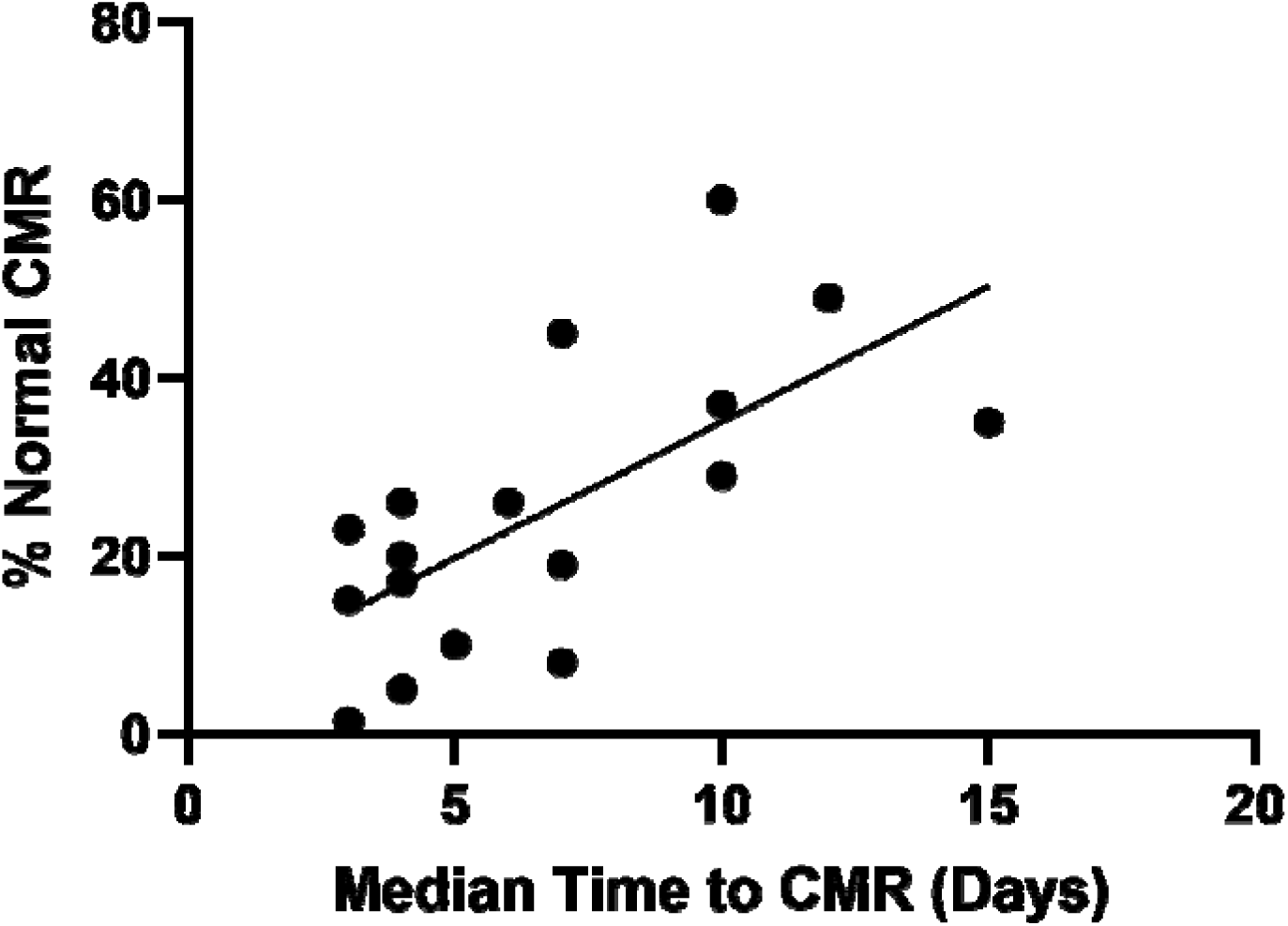
Correlation between time to CMR and a normal CMR study

**Figure 4.**
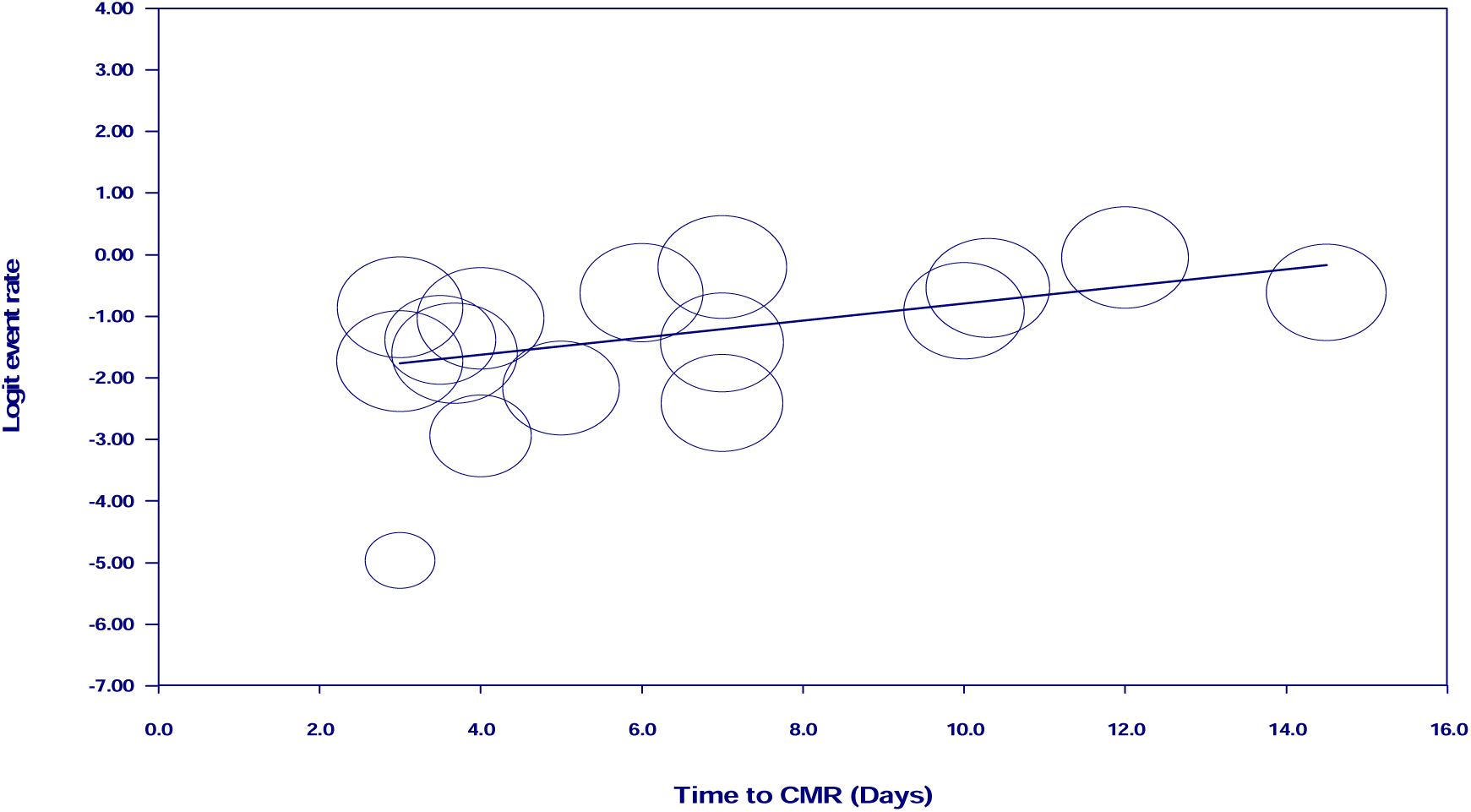
Meta-regression of time to CMR for normal CMR study.

## DISCUSSION

This meta-analysis provides evidence of a relationship between the time to CMR and diagnosis by CMR in patients with suspected MINOCA. The most common CMR diagnoses identified were myocarditis, myocardial infarction, Takotsubo syndrome, cardiomyopathy, and almost one-quarter had no cardiac diagnosis (normal CMR findings). There was a correlation between length of time to CMR and a normal CMR result (i.e., no cardiac diagnosis). Therefore, the longer the time it takes to have a CMR the greater the chance a patient will have no cardiac diagnosis on CMR. These findings highlight the clinical importance of using CMR and supports the use of CMR as early as possible in suspected MINOCA.

The findings of our study are similar to another study of 204 consecutive patients who presented with an ACS, troponin-positive and suspected of having MINOCA received a CMR scan (39). CMR timing was defined as “early” <2 weeks or “late” >2 weeks after presentation. There was a higher diagnostic yield by CMR among patients scanned early (84% vs. 57%, p<0.0001). Just over one-third of patients with myocarditis (33%) were diagnosed early compared to those with myocardial infarction who were diagnosed late (26%) (39). Moreover, patients with Takotsubo syndrome (p=0.002) and myocarditis (p=0.04) were more commonly represented in those patients who had an early CMR scan, whereas, there was no difference in rates of myocardial infarction based on scan time (p=1.00) (39). During this two-week time frame, CMR was able to provide a diagnosis for 70% of patients with suspected MINOCA (myocarditis 27%, myocardial infarction 26%, Takotsubo syndrome (9%), other cardiomyopathies 9%, and normal cardiac findings (30%) (39).

In addition, a recent study that assessed 255 troponin-positive acute chest pain and MINOCA patients who all had a CMR scan within seven days of coronary angiography (40). The patients were 42±16 years old, 65% male, and had a mean time to CMR of 2.7 days. During this one-week time frame, CMR was able to provide a diagnosis for 86% of MINOCA patients (myocarditis 53%, myocardial infarction 22%, Takotsubo syndrome (10%), myocardial contusion (0.39%), and normal heart (13%). Univariate analysis revealed various factors were associated with mortality (e.g., Takotsubo syndrome, myocardial infarction, age, hypertension, diabetes, female sex, ejection fraction, stroke volume index). However, the only independent predictors of mortality were hypertension and circumferential mechanical dispersion measured by strain analysis (40). This study highlighted the importance of establishing a correct diagnosis early using CMR and that this imaging modality resulted in patient lifestyle changes and closer surveillance.

In the current study, the median time to CMR scan was twelve and a half days with a range from zero (same day as presentation) to 365 days. These variations may be due to differences in healthcare systems, local clinical management protocols and/or guidelines, and availabilities in CMR services. Delays in providing optimal care may result in increases in cardiovascular morbidity and mortality (40).

In addition, a recent US analysis of Medicare data (between 2012-2017) reported that there had been a 75.5% increase in physician requests and use of CMR across 45 states (41), with higher rates of use by institutions from the Eastern states of the country. The current study identified that geographical location was also an important factor in terms of how quickly a patient received a CMR scan with this happening faster in the United States than in Europe, Australia, and lastly the United Kingdom. However, this data should be interpreted with caution because the information is gathered from research studies and not clinical service audits.

Imaging of the heart may involve echocardiography, CT, nuclear imaging modalities, and CMR. CMR has a number of advantages including its precision to assess anatomy and function, its safety (no ionizing radiation), high spatial resolution, 3D coverage, and excellent myocardial tissue characterisation capabilities (5). Of note, the current study did not assess how CMR relates to the use of other different diagnostic tests used at initial presentation. However, a recent study of 123 patients who had both echocardiography and CMR on the same day of presentation found that almost one-third (n=33) were classified as normal based on echocardiography findings (42). When these ‘normal’ patients had CMR, 70% were re-classified with a different diagnosis. This finding supports the use of CMR to diagnose these patients even after having other diagnostic tests with low troponin levels and normal echocardiography.

Nickander et al. (43) conducted a follow-up study of 113 MINOCA patients initially screened using CMR within 4 days of presentation and were invited to have a follow-up CMR scan at six-months. At follow-up, 65% of patients had a normal CMR scan and half of the patient had normalised the initial CMR diagnosis (43). This prospective study provides evidence for the early use of CMR and the benefit of late gadolinium enhancement (LGE) at 6-month follow-up to identify infarcts. Additional studies have also found that CMR provides a high (76%) diagnostic yield (44).

### Study Strengths and Limitations

The main strengths of this study include the comprehensive literature search strategy used with multiple databases to access all possible studies. We conducted a quality assessment of all studies and all studies were of high quality. A limitation of this study is the diagnostic criteria used to assess each condition using CMR, these were not always clearly defined, although most studies followed a clinical algorithm with selection criteria (45).

## Conclusions

This meta-analysis suggests that the longer it takes a patient to have a CMR for the assessment of suspected MINOCA, the more likely the result will be normal MR findings and no cardiac diagnosis made by CMR. We recommend that all suspected cases of MINOCA have a diagnostic CMR scan within the first ten days from presentation as the majority (>80%) of cases for all cardiac conditions are diagnosed within this period.

## Ethical Approval

This study is a meta-analysis of already published studies that have all been subject to ethical approval, and ethical approval for this meta-analysis is not applicable.

## Data Availability

All data produced in the present study are available upon reasonable request to the authors.

---

**Table 1.** Study characteristics for various cardiovascular outcomes.

| **Study** | **Region** | **Number of Patients** | **Mean Age** | **% Female** | **Median Time to CMR (days)** | **% Myocarditis** | **% TTS** | **% MI** | **% CM** | **% Normal CMR** |
| --- | --- | --- | --- | --- | --- | --- | --- | --- | --- | --- |
| Assomull *et al.* 2007 | Europe, UK | 60 | 44.0 | 28.0 | 14.5 median (1-90) | 50.0 | 1.7 | 11.6 | 1.7 | 35.0 |
| Laraudogoitia Zaldumbide *et al.* 2009 | Europe, Spain | 80 | 48.0 | 19.0 | mean 4+/-3 | 63.0 | 11.0 | 15.0 |  | 5.0 |
| Chopard *et al.* 2011 | Europe, France | 87 | 53.0 | 59.8 | 10.3 median (5-22) | 26.4 | 11.5 | 22.7 |  | 36.7 |
| Monney *et al.* 2011 | Europe, UK | 79 | 45.0 | 30.0 | 4 median (0-364); mean 33 | 81.0 | 6.0 | 13.0 |  |  |
| Leurent *et al.* 2011 | Europe, France | 107 | 43.5 | 38.0 | 5 median (mean 6.9 +/- 9) | 59.9 | 14.0 | 15.8 |  | 10.3 |
| Mahmoudi *et al.* 2012 | Europe, UK | 91 | 53.1 | 59.3 | 60 median (0-365) | 16.0 |  | 11.0 |  | 73.0 |
| Collste *et al.* 2012 | Europe, Sweden | 150 | 58.0 | 65.0 | 12 median (IQR 6-28) | 7.0 | 19.0 | 19.0 | 2.0 | 49.0 |
| Dastidar *et al.* 2019 | Europe, UK | 388 | 56.0 | 48.0 | 37 median | 25.0 |  | 25.0 | 25.0 | 26.0 |
| Bhatia *et al.* 2019 | North America, United States | 215 | 55.0 | 49.0 | 3.68 mean +/- 6 | 32.0 |  | 22.0 | 29.0 | 17.0 |
| Heidecker *et al.* 2019 | Europe, Switzerland | 556 | 57.0 | 29.5 | 10 median (IQR 4-23) | 13.7 | 1.3 | 7.4 | 8.8 | 60.0 |
| Emrich *et al.* 2021 | Europe, Germany | 145 | 58.0 | 33.0 | 3 median (IQR 1-5) | 33.1 | 9.7 | 15.1 | 40.7 | 1.38 |
| Gerbaud et al. 2020 | Europe, France | 40 | 50.2 | 37.5 | 3.5 median (IQR 2-8) |  |  | 77.5 |  | 20.0 |
| Hausvater et al. 2019 | Oceania, Australia<br>Europe, Sweden<br>North America, United States | 292 | 57.0 | 68.0 | 7 median (IQR 3-17) (R 1-121) | 13.0 | 14.0 | 19.0 | 9.0 | 45.0 |
| Lamour et al. 2020 | Europe, France | 135 | 48.0 | 33.4 | 7 median (IQR 3-14) | 62.0 | 4.4 | 14.1 |  | 19.3 |
| Lintingre et al. 2020 | Europe, France | 229 | 56.0 | 45.0 | 4 median (IQR 2-8) | 25.0 | 10.0 | 24.0 |  | 26.0 |
| Opolski et al. 2019 | Europe, Poland | 31 | 62.0 | 55.0 | 4 median (IQR 2-5) |  |  | 52 |  |  |
| Reynolds et al. 2021 | North America, United States | 116 | 58.0 | 100.0 | 6.0 median (IQR 3.5-9.0) | 14.7 | 3.4 | 32.8 | 2.6 | 25.9 |
| Sörensson et al. 2021 | Europe, Sweden | 148 | 58.0 | 71.0 | 3 median (IQR 2-3) | 17.0 | 35.0 | 22.0 | 3.0 | 23.0 |
| Tayal et al. 2020 | Europe, Denmark | 34 | 54.0 | 53.0 | 22 median (IQR 7-43) |  |  | 56.0 |  |  |
| Williams et al. 2022 | Europe, London | 719 | 57.0 | 51.0 | 30 median (6-54) | 26.0 | 12.0 | 26.0 | 10.0 | 26.0 |
| Yu et al. 2023 | Australia, Sydney | 70 | 52.8 | 44.0 | 10 median (4-54) | 21.0 | 9.0 | 15.0 | 8.0 | 29.0 |
| Konst et al. 2023 | North America, United States | 252 | 54 | 57 | 3 median (2-8) | 13.0 |  | 25.0 | 44.0 | 15.0 |
| Junca et al. 2024 | Europe,<br>Spain | 207 | 50 | 40 | 7 median (4-12) | 45.0 | 19.0 | 20.0 | 7.0 | 8.0 |

TTS: Takotsubo syndrome; MI: Myocardial infarction; CM: Cardiomyopathy.

**Supplementary Table 1.** Quality assessment results.

| Study | Inclusion Criteria | Measurement | Valid Methods | Consecutive sample | Complete Inclusion | Demographics | Clinical Information | Outcomes | Generalizability | Statistics | Score |
| --- | --- | --- | --- | --- | --- | --- | --- | --- | --- | --- | --- |
| Assomull et al. 2007 | ✓ | ✓ | ✓ | ✓ | ✓ | ✓ | ✓ | ✓ | ✓ | ✓ | 10 |
| Laraudogoitia Zaldumbide et al. 2009 | ✓ | ✓ | ✓ | 0 | ✓ | ✓ | ✓ | ✓ | ✓ | 0 | 8 |
| Chopard et al. 2011 | ✓ | ✓ | ✓ | ✓ | ✓ | ✓ | ✓ | ✓ | ✓ | ✓ | 10 |
| Monney et al. 2011 | ✓ | ✓ | ✓ | 0 | ✓ | ✓ | ✓ | ✓ | ✓ | ✓ | 9 |
| Leurent et al. 2011 | ✓ | ✓ | ✓ | ✓ | ✓ | ✓ | ✓ | ✓ | ✓ | ✓ | 10 |
| Mahmoudi et al. 2012 | ✓ | ✓ | ✓ | ✓ | ✓ | ✓ | ✓ | 0 | 0 | ✓ | 8 |
| Collste et al. 2012 | ✓ | ✓ | ✓ | ✓ | 0 | ✓ | ✓ | ✓ | ✓ | ✓ | 9 |
| Dastidar et al. 2019 | ✓ | ✓ | ✓ | ✓ | ✓ | ✓ | ✓ | ✓ | ✓ | ✓ | 10 |
| Bhatia et al. 2019 | ✓ | ✓ | ✓ | 0 | ✓ | ✓ | ✓ | ✓ | ✓ | ✓ | 9 |
| Bettina et al. 2019 | ✓ | ✓ | ✓ | 0 | ✓ | ✓ | ✓ | ✓ | ✓ | ✓ | 9 |
| Emrich et al. 2021 | ✓ | ✓ | ✓ | 0 | 0 | ✓ | ✓ | ✓ | ✓ | ✓ | 8 |
| Gerbaud et al. 2020 | ✓ | ✓ | ✓ | ✓ | ✓ | ✓ | ✓ | ✓ | ✓ | ✓ | 10 |
| Hausvater et al. 2019 | ✓ | ✓ | ✓ | 0 | ✓ | ✓ | ✓ | ✓ | ✓ | ✓ | 9 |
| Lamour et al. 2020 | ✓ | ✓ | ✓ | 0 | ✓ | ✓ | ✓ | ✓ | ✓ | ✓ | 9 |
| Lintingre et al. 2020 | ✓ | ✓ | ✓ | ✓ | ✓ | ✓ | ✓ | ✓ | ✓ | ✓ | 10 |
| Opolski et al. 2019 | ✓ | ✓ | ✓ | ✓ | ✓ | ✓ | ✓ | ✓ | ✓ | ✓ | 10 |
| Reynolds et al. 2021 | ✓ | ✓ | ✓ | 0 | ✓ | ✓ | ✓ | ✓ | ✓ | ✓ | 9 |
| Tayal et al. 2020 | ✓ | ✓ | ✓ | 0 | ✓ | ✓ | ✓ | ✓ | ✓ | ✓ | 9 |
| Sörensson et al. 2021 | ✓ | ✓ | ✓ | ✓ | ✓ | ✓ | ✓ | ✓ | ✓ | ✓ | 10 |
| Willaims et al. 2022 | ✓ | ✓ | ✓ | ✓ | ✓ | ✓ | ✓ | ✓ | ✓ | ✓ | 10 |
| Yu et al. 2023 | ✓ | ✓ | ✓ | ✓ | ✓ | ✓ | ✓ | ✓ | ✓ | ✓ | 10 |
| Konst et al. 2023 | ✓ | ✓ | ✓ | ✓ | ✓ | ✓ | ✓ | ✓ | ✓ | ✓ | 10 |
| Junca et al. 2024 | ✓ | ✓ | ✓ | 0 | ✓ | ✓ | ✓ | ✓ | ✓ | ✓ | 9 |

✓: Yes; 0: No or unclear

